# Objective adherence to an online FAVAS therapeutic game for treating amblyopia in children

**DOI:** 10.1101/2021.01.27.21250294

**Authors:** Catheline Bocqué, Jingyun Wang, Annekatrin Rickmann, Henrike Julich-Haertel, Uwe Kaempf, Kai Januschowski

**Affiliations:** Klaus Heimann Eye Research Institute, An der Klinik 10, 66280 Sulzbach, Germany; SUNY College of Optometry, 33 W 42nd St, New York, NY 10036, United States; Caterna Vision GmbH, David-Gilly-Str. 1, 14196 Potsdam, Germany; Centre for Ophthalmology, University Eye Hospital Tuebingen, Schleichstr. 12, 72076 Tuebingen, Germany

## Abstract

**Aim:** This retrospective study was to evaluate whether an updated version of attention binding digital therapeutic games based on the principle of Focal Ambient Visual Acuity Stimulation (FAVAS) would result in an improved patient adherence of patching in 4- to 12-year-old patients with amblyopia.

**Methods:** We analyzed pseudonymised electronically recorded data from patients treated with two different versions of attention binding digital therapeutic games in 2015 and 2020. Two groups of children treated with occlusion therapy and attention binding digital therapeutic games, divided in treatment version, were compared. Patients in Group 2015 used the old version of therapeutic games without tablet computer functionality, while Group 2020 used more attractive therapeutic games with tablet computer functionality. Objective adherence was calculated by comparing the amount of minutes using the therapeutic games as monitored in the automatized logbook versus prescribed minutes of using the games.

**Results:** Children in Group 2015 spent on average 2009.3±1372.1 (36 to 5472) minutes using FAVAS; children in Group 2020 spent on average 2695.5±1526.8 (37.5 to 5672) minutes using the improved therapy. Meaning, Group 2020 spent 686.2 more minutes on FAVAS than Group 2015 (t=3.87, P<0.001). Although patient adherence was very variable, it significantly improved up to 78% ± 46% in Group 2020 compared to the 57% ± 34% in Group 2015 (t=4.3, P<0.001).

**Conclusion:** FAVAS 2020 with an improved gamification aspect as well as tablet computer functionality increased adherence significantly compared to the earlier version FAVAS 2015, indicating that FAVAS 2020 could be an effective approach to support patching amblyopia treatment.

## Introduction

Amblyopia is a developmental disorder resulting in decreased visual acuity in one eye. Decreased stimulation by the weaker eye during the developmental phase of vision leads to underdevelopment of the corresponding cortical visual areas, making the eye amblyopic.^1^ Amblyopia is one of the most common ophthalmological disorders in children and has a lasting effect on the individuals’ quality of life.^2^ While affected children are impacted in their daily activities and future job selection, it also increases the risk of a severe trauma for the better fellow eye.^3^ Since Sattler (1927) occlusion therapy, after binocular eyeglass correction, has been the standard therapeutic approach forcing visual development of the affected weak eye by an input deprivation of the better seeing eye.^4^ However, by applying this therapy, a high rate of patients (approximately 25% to 30%) do not show a full recovery of visual function and some of those patients even show further worsening in visual function.^5-8^ Visual acuity improvement in the amblyopic eye is significantly impacted by adherence of the patching therapy (Al-Zuhaibi S et al. 2009).^9^ For a long time, a system of monocular and binocular visual exercises and stimulation methods (pleoptics and orthoptics) in support of the standard occlusion treatment has been developed (Otto & Rabethge 1963; Otto & Stangler 1969), but only with limited success.^11-13^ To improve the adherence of the occlusion therapy, a gamification of therapy could be helpful and has been implemented in several approaches. Monocular Focal Ambient Visual Acuity Stimulation (FAVAS) therapeutic games are an innovative digital therapeutic games approach designed as a supplementary treatment to patching. A customized moving ambient sinusoidal wave pattern (moving gratings) is presented in the background of focal attention binding digital therapeutic games, stimulating cortical areas to activate the central perceptive activity of the amblyopic eye again and thus improving visual acuity.^14^ FAVAS differs in several ways from the moving grating stimulation Cambridge Stimulator (CAM) treatment. CAM used high-contrast square-wave gratings, which were rotated in front of the amblyopic eye while playing on a transparent cover in front of the stimulator. It was initially reported to improve outcome combined with patching, but failed to succeed in subsequent prospective randomized controlled studies.^15,16^ Beyond CAM treatment, FAVAS relies not only on spatial frequency selectivity of the ambient background stimulus, but also on an interaction of its coordinated temporal frequency parameters with the focal sensory-motoric gaming activity (Kämpf et al. 2008).^14,17,18^ Previously, Kämpf et al. showed that FAVAS had a promising effect. Since the early version of FAVAS focused on the therapeutic aspect of visual acuity stimulation and put less focus on the user-friendliness, gamification and attention binding aspect, this modification could potentially impact patient adherence. Apart from other technical updates, later versions of commercially available treatment games specifically improved this aspect.

The goal of this study is to evaluate whether an improved gamification aspect as well as tablet computer functionality of FAVAS therapeutic games would result in a higher patient adherence compared to the earlier version. Therefore, we analyzed the electronically recorded data from a commercially available FAVAS system (Caterna Vision GmbH, Potsdam, Germany) in 4- to 12-year-old patients, and compared adherence to the earlier version FAVAS 2015 with adherence to an updated version FAVAS 2020.

## Methods

This retrospective study adhered to the declaration of Helsinki, was conducted at a single center and was approved by the local ethics committee (118/19, trial registration DRKS00017633). Due to the retrospective nature of this study and an pseudonymisation at the source, no additional informed consent was required.

We compared pseudonymised electronic user protocols showing the therapeutic game activity time of patients aged between 4 and 12 years; all patients were diagnosed with amblyopia by their ophthalmologist, treated with a combination of occlusion and FAVAS therapy (Caterna Vision GmbH, Potsdam, Germany). The amblyopia was associated with anisometropia and/or mild strabismus. Patients had their current refractive correction worn for at least 16 weeks until two consecutive visual acuity measurements, at least eight weeks apart, did not change by more than 1 logMAR line. The amblyopic eye had a best-corrected visual acuity (BCVA) from 20/40 to 20/200; the fellow eye a BCVA of 20/32 or better, and the difference between the eyes was ≥ 3 logMAR lines. We excluded children with proven learning disabilities, known epilepsy, or with other pre-existing ophthalmic conditions or deprivation amblyopia (weak vision duo to an organic cause).

### Treatments

All patients had full binocular correction with glasses, which were prescribed by their local eye doctors. For occlusion therapy patients used standard eye patches like opticlude (3M), Piratoplast etc. Every individual got a personalized occlusion rhythm of how many hours per day they had to wear the patch, depending on the visual acuity, fixation site at the fundus, age and other findings. Each participant was provided with an access to a home-based FAVAS, offered by the Caterna Vision GmbH. The prescribed FAVAS games therapy was 90 days long, played every day for 30-45 minutes during occlusion time. The treatment was reimbursed for the home-based stimulation therapy by their insurance company.

Group 2015 (n=138) contains a dataset of patients which used FAVAS version 1.0 in 2015. They had to read instructions for the games. For playing therapeutic games, only keyboard and mouse at a fixed screen size of 15 inches were available. Group 2020 (n=129) contains a dataset of patients with therapy in 2020. They were able to play directly with high resolution graphics and high usability. For playing games, not only keyboard and mouse but also touchscreen at screens between 10 to 27 inches were available.

### Modification of attention binding online games

The FAVAS 1.0 therapy was modified in a few ways: In terms of technical refinement, a larger selection with a variety of engaging games to attract children’s attention and participation was included, resulting in nine edutainment HTML5 games for children between 4 to 12 years. There was a backward compatibility for browser, screen size and hardware combined with better onboarding (patient manual, AQ, simplified usability). The majority of children between 4 to 12 years has access to a tablet computer which makes the access to the therapy independent from time and place. Therefore we focused to make the therapy effective on tablet computers.

### Main outcome measures

The primary outcome measure was the therapy adherence. Objective adherence was defined by comparing the amount of minutes using the computer game as monitored in the automatized logbook versus prescribed minutes of using the game.

### Statistical analysis

Sample size estimates were based on data from literature reviews and data from participants in Group 2015 pilot trials who would meet the eligibility criteria for the current protocol.^2,3,5,6,13^ With our sample size of Group 2020, the effect size between the 2 groups is 0.51. Analysis of variances were used to analyze group differences in continuous variables. The Pearson’s Chi-squared test was used for analyzing categorical variables. P-values smaller than 0.05 were considered statistically significant.

## Results

In Group 2015, a total of 138 patients were analyzed; in Group 2020, a total of 129 patients were analyzed. Basic characteristics of the two groups, such as age, sex and types of amblyopia are shown in Table 1. The mean age was slightly younger in Group 2020 than the Group 2015, by approximately 0.6 year. Both groups had similar gender ratios and amblyopia types distribution.

**Table 1.** Baseline characteristics

|  | Group 2015<br>*n*=138 | Group 2020<br>*n*=129 | *P*-value |
| --- | --- | --- | --- |
| Sex |  |  |  |
| Female no. (%) | 70 (50.7) | 67 (51.9) | Chi-squared=0.005,<br>P=0.93 |
| Male no. (%) | 68 (49.3) | 62 (48.1) |  |
| Age (years) |  |  |  |
| Mean±SD (min, max) | 7.8±2.1<br>(4.2, 15.6) | 7.2±2.3<br>(4.3, 14.7) | t=2.22, P=0.03* |
| Amblyopia Types |  |  |  |
| Strabismic/Anisometropic | 65/73 | 61/68 | Chi-squared=0,<br>P=1 |

### Adherence

Children in Group 2015 spent on average 2009.3±1372.1 (36 to 5472) minutes of time on FAVAS games; children in Group 2020 spent on average 2695.5±1526.8 (37.5 to 5672) minutes on playing. Meaning that Group 2020 spent on average 686.2 minutes more time on FAVAS games than Group 2015 (t=3.87, P<0.001). In both groups, some patients had a high adherence of more than 150%, which indicates that some individuals enjoyed FAVAS treatment (Figure 2). In Group 2015 the mean adherence was 57% ± 34% of the prescribed exercise time. Adherence in Group 2020 significantly improved up to 78% ± 46% compared to Group 2015 (t=4.3, P<0.001).

**Figure 1:**
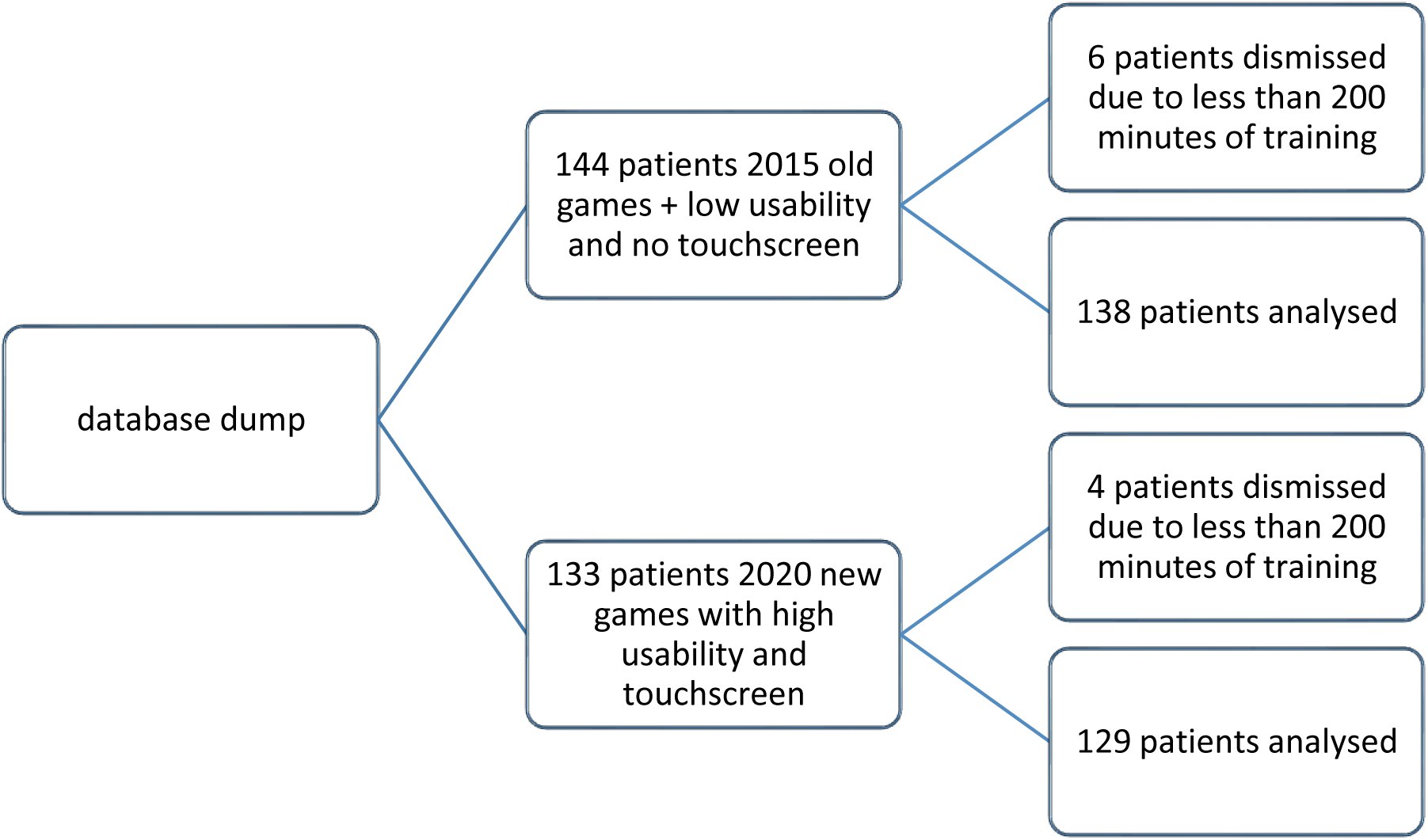
Patient flow chart with Group 2015 having six dropouts and Group 2020 four dropouts due to technical challenges.

**Figure 2.**
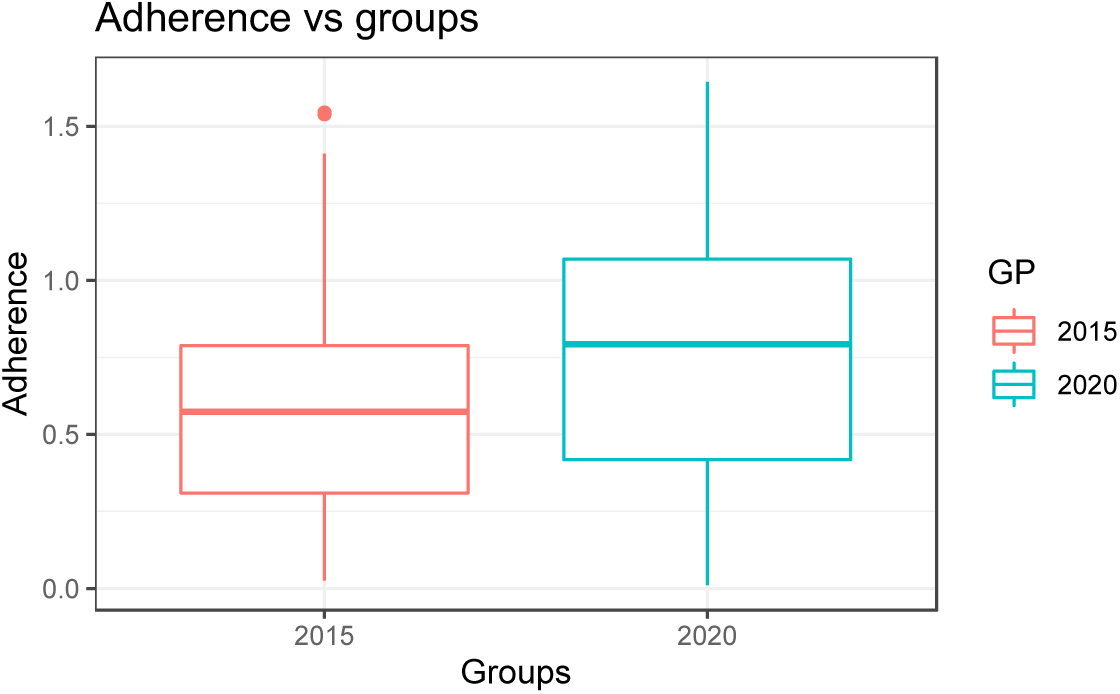
Boxplot of adherence with FAVAS treatment games in Group 2015 and Group 2020.

## Discussion

In this study we could show that improving the gaming aspect and usability of FAVAS therapy can enhance patient adherence, thus possibly improve overall therapeutic effect. In light of the actual pandemic but also considering problematic access to specialists these data are encouraging and give us important information about the acceptance of online treatment strategies. This suggests that FAVAS with a larger selection of games to attract active participation, backward compatibility for browsers, screen size and hardware combined with better onboarding (patient manual, FAQ, simplified usability) is a good strategy to improve patient adherence. These findings are supported by other studies showing that common interventions (Cartoons, education, Sticker-Games) have been effective to improve compliance.^19^

Considering occlusion therapy it was shown in earlier studies that adherence is rather low with only 60% adherence.^2,3,5^ Our adherence was better (80%) during the FAVAS therapy indicating that this might be beneficial for overall treatment adherence, especially for patients with low motivation for occlusion as monotherapy. However, our data do not give a precise information about the rest of the occlusion time therefore this conclusion should be regarded critically; they should be evaluated further regarding functional outcome such as visual acuity or contrast sensitivity improvement in further studies.^20^ An overview about therapy adherence studies is shown in Table 2 and an average of occlusion compliance in Figure 3.

**Table 2.**
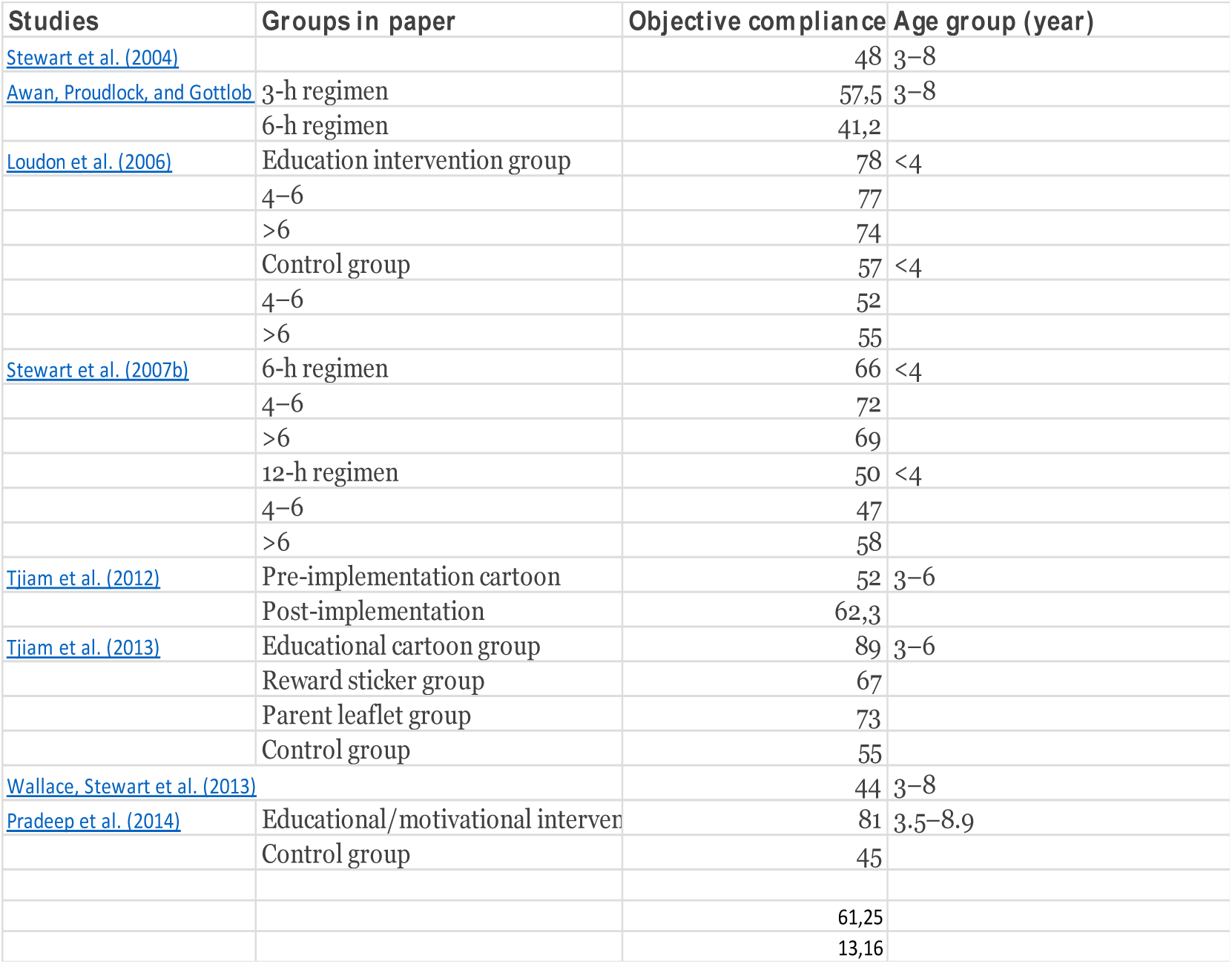
Literature overview of 24 occlusion compliances with standard regimens to interventions like education, cartoons, stickers to boost motivation in children and parents.^10^

**Figure 3.**
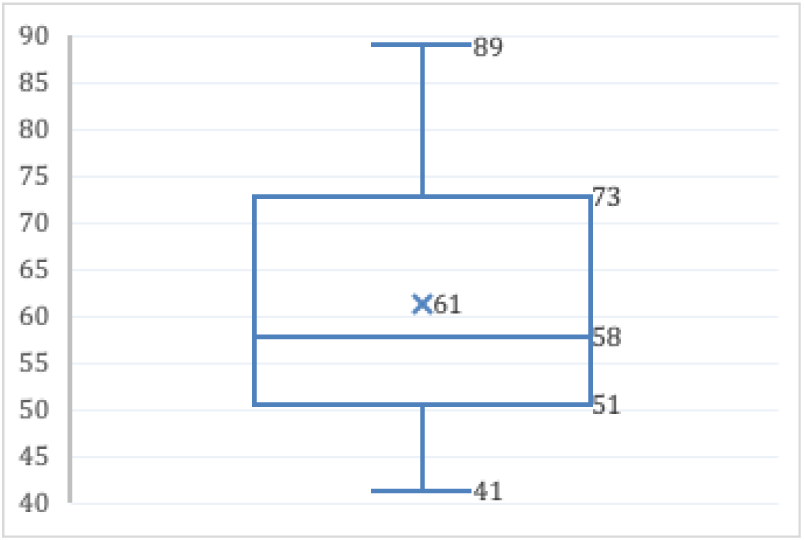
Boxplot of average occlusion compliance of meta-analysis of 24 studies from Wang et al. 2015.^10^

Our study has a few limitations: the retrospective character and only self-reported data about patching should be regarded critically. During ongoing therapeutic interventions, the adherence to patching treatment is on average continuously decreasing as the longer the treatment lasts.^10^ Thus, our future tasks will not only be increasing average adherence, but also to change the current dynamic of adherence by slowing, maintaining or reversing the decreasing adherence trend with computer-assisted therapeutic interventions. The data of this study show an attractive option: improving gamification and add tablet computer functionality might stop this negative dynamic, however further studies with automatic occlusion dose monitoring should be added to verify our results.

## Data Availability

we analyzed the electronically recorded data from a commercially available FAVAS system (Caterna Vision GmbH, Potsdam, Germany)

## Notes

### Competing Interest Statement

The authors have declared no competing interest.

### Clinical Trial

DRKS00017633

### Funding Statement

-

### Author Declarations

Ethics committee: Arztekammer des Saarlandes, Korperschaft des offentlichen Rechts, Ethikkommission. Ethical approval was given on 27.05.2019. Nr.118/19

## References

1. Hubel DH, Wiesel TN. The period of susceptibility to the physiological effects of unilateral eye closure in kittens. J Physiol. 1970;206(2):419–436.

2. Elflein HM, Fresenius S, Lamparter J, et al. The Prevalence of Amblyopia in Germany: Data from the Prospective, Population-Based Gutenberg Health Study. Dtsch Arztebl Int 2015;112:338–44.

3. Maurer D, Mc KS. Classification and Diversity of Amblyopia. Vis Neurosci 2018;35:E012.

4. Loudon SE, Simonsz HJ. The History of the Treatment of Amblyopia, Strabismus 2005, Volume 13; 93–106.

5. Repka MX, Beck RW, Holmes JM, et al. A Randomized Trial of Patching Regimens for Treatment of Moderate Amblyopia in Children. Arch Ophthalmol 2003;121:603–11.

6. Repka MX, Cotter SA, Beck RW, et al. A Randomized Trial of Atropine Regimens for Treatment of Moderate Amblyopia in Children. Ophthalmology 2004;111:2076–85.

7. Repka MX, Wallace DK, Beck RW, et al. Two-Year Follow-up of a 6-Month Randomized Trial of Atropine Vs Patching for Treatment of Moderate Amblyopia in Children. Arch Ophthalmol 2005;123:149–57.

8. Wallace DK, Pediatric Eye Disease Investigator G, Edwards AR, et al. A Randomized Trial to Evaluate 2 Hours of Daily Patching for Strabismic and Anisometropic Amblyopia in Children. Ophthalmology 2006;113:904–12.

9. Al-Zuhaibi S, Al-Harthi I, Cooymans P, Al-Busaidi A, Al-Farsi Y, Ganesh A. Compliance of amblyopic patients with occlusion therapy: A pilot study. Oman J Ophthalmol. 2009 May;2(2):67–72. doi:10.4103/0974-620X.53035. PMID: 20671832; PMCID: PMC2905182.

10. Wang J. Compliance and Patching and Atropine Amblyopia Treatments. Vision Res 2015;114:31–40.

11. Otto J, Rabetge G. Pleoptische Schulung und auftreten des ‘Wendephaenomens’ [pleoptic training and the appearance of the ‘version phenomen’]. Klin Monbl Augenheilkd. 1963 Nov;143:524–9. German. PMID: 14097581.

12. Otto J, Stangler E. Wirkung optomotorischer Reize auf Fixationsort und Auflösungsvermogen amblyoper Augen mit exzentrischer Fixation [Effect of optomotor stimulation on the kind of fixation and correction of amblyopic eyes with excentric fixation]. Ophthalmologica. 1969;157(2):135–41. German. doi:10.1159/000305637. PMID: 5790897.

13. Bangerter A. [Amblyopia Therapy]. Bibl Ophthalmol 1953;112:1–96

14. Kämpf U, Shamshinova A, Kaschtschenko T, Mascolus W, Pillunat L, Haase W. Long-term application of computer-based pleoptics in home therapy: selected results of a pro-spective multicenter study. Strabismus. 2008;16(4):149–58. doi:10.1080/09273970802451125. PMID: 19089760.

15. Campbell FW, Hess RF, Watson PG, Banks R. Preliminary results of a physiologically based treatment of amblyopia. Br J Ophthalmol. 1978;62:748–55.)

16. Lennerstrand G, Samuelsson B. Amblyopia in 4-Year-Old Children Treated with Grating Stimulation and Full-Time Occlusion; a Comparative Study. Br J Ophthalmol 1983;67:181–90.

17. Kämpf U, Muchamedjarow F, Seiler T. Unterstützende Amblyopiebehandlung durch Computerspiele mit Hintergrundstimulation: Eine 10-tägige plazebokontrollierte Pilot-Studie [Supportive amblyopia treatment by means of computer games with background stimulation; a placebo controlled pilot study of 10 days]. Klin Monbl Augenheilkd. 2001 Apr;218(4):243–50. German. doi:10.1055/s-2001-14921. PMID: 11392270.

18. Kämpf U, Rychkova S, Muchamedjarow F, Heim, E. Comparing the results of the application of moving and stationary sinusoidal gratings in the functionally assisted treatment of meridional amblyopia. Poster session presented at the European Conference on Visual Perception 2017, Berlin, Germany. Retrieved from URL: http://journals.sagepub.com/page/pec/collections/ecvp-abstracts/index/ecvp-2017 retrieved on 2020-09-29

19. Tjiam AM, Holtslag G, Van Minderhout HM, et al. Randomised Comparison of Three Tools for Improving Compliance with Occlusion Therapy: An Educational Cartoon Story, a Reward Calendar, and an Information Leaflet for Parents. Graefes Arch Clin Exp Ophthalmol 2013;251:321–9.

20. Januschowski K, Bechtold TE, Schott TC, et al. Measuring Wearing Times of Glasses and Ocular Patches Using a Thermosensor Device from Orthodontics. Acta Ophthalmol 2013;91:e635–40.

